# Correlating Lignin Positivity with Self-Reported Administration of Sulfadoxine-Pyrimethamine (SRA-Sp) among Pregnant Women

**DOI:** 10.1101/2025.08.01.25332604

**Authors:** Dhikrullah A. Adebayo, Waheed A. Adedeji, Aminat Alarape-Raji, Sulayman Tunde Balogun, Bukola Adeola Adaramola, Fatai A. Fehintola

## Abstract

**Background:** This study explores the relationship between lignin positivity and self-reported sulfadoxine-pyrimethamine (SP) administration in pregnant women residing in malaria-endemic regions of Yemetu, Ibadan. Sulfadoxine-pyrimethamine, a commonly used malaria prevention drug during pregnancy, has shown varying levels of efficacy based on individual adherence to treatment regimens. Lignin positivity, a potential biomarker associated with oxidative stress, was assessed as a correlate of therapeutic response to SP and its potential role in monitoring adherence.

**Materials and Methods:** The study employed a longitudinal design involving 355 pregnant women attending Adeoyo Maternity Teaching Hospital, Yemetu, Ibadan, aged 18-45, who had received SP for malaria prophylaxis. Self-reported data on SP administration were collected through a researcher’s self-structured questionnaire, while lignin positivity was determined through biochemical lignin tests on the urine of the pregnant women conducted at three intervals (Day 0, Day 7, and Day 28).

**Results:** The results demonstrated a significant increase in lignin positivity over the 28-day period (Day 0 to Day 7 (p > 0.000) at 91.3% increase, and 7 to Day 28 (p > 0.007) with 93.5% rate of lignin positivity rise), with the highest positivity observed at Day 28. This trend was statistically significant, indicating a strong correlation between SP administration and increased lignin test results.

**Conclusion:** There existed strong evidence supporting a statistically significant correlation between lignin positivity and the self-reported administration of sulfadoxine-pyrimethamine (SP) among pregnant women attending Adeoyo Maternity Teaching Hospital, Yemetu, Ibadan. This implies that lignin positivity is a useful potential biomarker for ascertaining SP adherence in pregnant women. These results highlight the effectiveness of sulfadoxine-pyrimethamine in malaria prevention and suggest that lignin positivity could serve as a valuable tool for assessing adherence to malaria chemoprophylaxis among pregnant women.

## 1. Introduction

Lignin, is one of such potential biomarkers and a complex polymer primarily found in plant cell walls. While lignin is well-known for its structural role in plants, emerging research suggests that lignin and its metabolites may be involved in oxidative stress processes in humans. Oxidative stress, characterized by an imbalance between reactive oxygen species (ROS) and antioxidants, plays a pivotal role in a variety of physiological and pathological conditions, including immune responses and drug metabolism [1]. Given the critical role that oxidative stress can have on drug metabolism and the immune system, lignin positivity could offer valuable insights into how the body, especially during pregnancy, responds to external factors like drug administration [2,3].

The presence of lignin or its metabolites in the human body, particularly in pregnant women, may serve as an indicator of metabolic changes that influence how drugs like sulfadoxine-pyrimethamine are processed. Recent studies [4,5,6] have suggested that elevated oxidative stress in pregnancy may affect the activity of cytochrome P450 enzymes, which are responsible for metabolizing various drugs, including SP. Therefore, understanding how lignin positivity correlates with the self-reported administration of SP among pregnant women could provide valuable information about the pharmacokinetics of SP, potentially leading to more effective and personalized treatment strategies.

As the relationship between lignin and oxidative stress remains an emerging area of research, it is essential to investigate whether lignin positivity is associated with the self-reported administration of IPTp-SP. A better understanding of this relationship could have significant implications for improving malaria treatment outcomes in pregnancy, optimizing drug dosing, and ensuring that pregnant women in malaria-endemic regions receive the full benefit of preventive treatments [7,8,9]. Exploring this link could also provide a deeper understanding of the mechanisms through which oxidative stress influences drug metabolism, paving the way for improved therapeutic strategies that consider the unique physiological changes occurring during pregnancy.

Sulfadoxine-pyrimethamine (SP) is a combination antimalarial drug widely used for the prevention and treatment of malaria, particularly in pregnant women. This drug is integral in malaria control strategies in regions where the disease is endemic, as it helps to reduce the incidence of malaria-related complications during pregnancy. Malaria in pregnancy poses significant risks, including maternal anemia, fetal growth restriction, preterm birth, and even maternal and neonatal mortality [10]. Therefore, SP is often administered as part of intermittent preventive treatment to protect both the mother and the fetus from these adverse outcomes. The drug works by inhibiting the folate synthesis pathway in the malaria parasite, thus preventing its replication and reducing the parasite load in the body.

Lignin, a complex polymer found in the cell walls of plants, has been recognized primarily for its structural role in plants. However, emerging research suggests that lignin and its metabolites may also serve as indicators of certain metabolic processes or oxidative stress within the human body [11,12]. Oxidative stress, characterized by an imbalance between reactive oxygen species and antioxidants, is a critical factor in many physiological and pathological processes, including immune responses and drug metabolism. The presence of lignin or its derivatives in the human body, particularly in pregnant women, has not been fully explored, though it may offer valuable insights into how the body responds to external factors such as drug administration. Lignin positivity, therefore, could reflect changes in metabolic or immune functions that occur during pregnancy, potentially influencing the way drugs like sulfadoxine-pyrimethamine are processed or utilized in the body [13,14].

The significance of investigating the correlation between lignin positivity and self-reported sulfadoxine-pyrimethamine administration lies in its potential to uncover biomarkers that could provide a clearer understanding of drug adherence and efficacy. Adherence to malaria prevention treatments like SP is critical for ensuring the drug’s effectiveness in reducing malaria-related risks during pregnancy [15]. However, self-reported data on drug administration can often be unreliable due to recall bias or underreporting. Identifying biomarkers, such as lignin positivity, could serve as an objective method to verify adherence and evaluate the body’s response to the drug. By correlating lignin positivity with SP administration, this research could open new avenues for improving maternal and fetal health outcomes in malaria-endemic regions, where accurate monitoring of drug adherence and treatment efficacy is vital. Moreover, the discovery of such biomarkers could lead to more personalized and effective approaches to malaria prevention in pregnancy, ultimately contributing to better health outcomes for both mothers and their babies.

Despite the widespread use of sulfadoxine-pyrimethamine (SP) for malaria prophylaxis in pregnancy, there remains a significant gap in understanding how adherence to treatment and the response to the drug can be accurately monitored. SP has proven to be an effective intervention for preventing malaria-related complications in pregnant women, but its full efficacy depends heavily on consistent and correct usage. However, challenges such as incomplete adherence, underreporting of medication intake, and variability in drug metabolism across different individuals can undermine its effectiveness. While self-reported adherence data provides some insights, it is often unreliable due to recall bias or social desirability bias, making it difficult to evaluate whether women are following the prescribed regimen.

There is a notable lack of biomarkers that can objectively predict adherence or assess the body’s response to SP treatment. Lignin positivity, a potential biomarker, may present a novel solution to this issue. Lignin, which is typically associated with plant structures, may indicate metabolic alterations or oxidative stress in the body, potentially reflecting the physiological impact of drug administration [9,8,10]. If lignin positivity correlates with self-reported SP usage, it could serve as an effective tool for monitoring adherence to malaria prevention in pregnant women. This would allow healthcare providers to more accurately assess whether women are taking the prescribed medications and how their bodies are responding to the drug. As such, lignin positivity could provide an important new approach to understanding the therapeutic journey of pregnant women undergoing malaria prophylaxis, helping to ensure that the treatment is both taken as prescribed and effective in preventing malaria-related complications. The researcher intended to investigate if there exist a correlation between lignin positivity and the self-reported administration of sulfadoxine-pyrimethamine among pregnant women.

H_o1_: There is no statistically significant correlation between lignin positivity and the self-reported administration of sulfadoxine-pyrimethamine (SP) among pregnant women.

H_1_: There is statistically significant correlation between lignin positivity and the self-reported administration of sulfadoxine-pyrimethamine (SP) among pregnant women.

### Sulfadoxine-Pyrimethamine and Malaria Prevention

Sulfadoxine-pyrimethamine (SP) has long been used as an effective preventive treatment for malaria, especially in pregnant women living in malaria-endemic regions. Malaria during pregnancy poses a significant threat to both maternal and fetal health, leading to complications such as maternal anemia, preterm birth, intrauterine growth restriction, stillbirth, and even maternal death [3,7,9]. SP, a combination of sulfadoxine (a sulfonamide) and pyrimethamine (a dihydrofolate reductase inhibitor), works by inhibiting the folate pathway in the Plasmodium parasite, thus preventing the parasite from multiplying in the body. This antimalarial drug is typically administered as intermittent preventive treatment (IPTp) during pregnancy, with the aim of reducing the malaria parasite load and preventing its associated risks [10].

The effectiveness of SP in preventing malaria during pregnancy is well-documented, with studies showing that it can significantly reduce the incidence of maternal malaria, low birth weight, and fetal complications when administered appropriately. However, its success is dependent on several factors, including correct dosage, timely administration, and adequate adherence to the prescribed regimen [11]. In regions where malaria transmission is high, SP has been a cornerstone of malaria prevention programs for pregnant women, with national policies advocating for its use as part of routine antenatal care.

Despite its widespread use, there are notable challenges that affect the effectiveness of SP in preventing malaria during pregnancy. One of the most significant challenges is poor adherence to the treatment regimen [16]. Adherence to SP can be influenced by various factors, such as the timing of administration, access to healthcare, cultural beliefs, and a lack of understanding about the importance of completing the full course of treatment [5,17]. Pregnant women in malaria-endemic regions may also face logistical barriers such as long distances to healthcare facilities, cost-related issues, and limited access to healthcare professionals who can provide guidance on proper medication use. These barriers often lead to inconsistent use of SP, reducing its overall efficacy [2,6,9].

In addition to logistical and behavioral factors, socio-economic influences play a critical role in the uptake of SP. Women from lower socio-economic backgrounds may face greater difficulties in accessing healthcare, which could contribute to lower adherence rates [12,14,18]. Socio-economic disparities can also affect a woman’s understanding of the importance of malaria prevention during pregnancy, with some women possibly undervaluing the need for regular IPTp due to competing priorities or a lack of education. Furthermore, misconceptions and cultural factors surrounding medication use during pregnancy can contribute to non-adherence or discontinuation of treatment, even when it is recommended by healthcare providers.

Given these challenges, improving adherence to SP and ensuring its optimal use during pregnancy remains a significant public health goal [13]. Efforts to enhance adherence include better patient education, improved access to healthcare services, and strategies to address socio-economic barriers. However, more research is needed to identify objective markers that could help assess adherence to treatment and understand the biological responses that may influence drug effectiveness in pregnant women [16,18]. Identifying such biomarkers could provide insights into improving treatment regimens, personalizing care, and ultimately enhancing maternal and fetal health outcomes.

### Lignin as a Biomarker

Lignin, a complex polyphenolic compound found in the cell walls of plants, is primarily known for its structural role in plant biology. However, over the years, there has been growing interest in exploring lignin and its derivatives as potential biomarkers for various health conditions in humans [6]. Recent researches has begun to suggest that lignin, or compounds derived from it, may be involved in several physiological processes, particularly those related to oxidative stress and immune response [5,6,9]. These properties have led to investigations into its potential as a biomarker for monitoring health conditions such as metabolic disorders, inflammation, and even responses to certain drugs.

While the use of lignin as a biomarker in human health remains relatively under-explored, some studies have proposed that lignin-related compounds might play a role in reflecting oxidative stress levels, which can be a critical factor in numerous diseases, including pregnancy-related conditions [15,17,19]. Oxidative stress occurs when there is an imbalance between reactive oxygen species (ROS) and antioxidant defenses, and it has been implicated in a variety of pregnancy complications, including preeclampsia, gestational diabetes, and fetal growth restriction. Lignin or its metabolites may serve as indicators of such oxidative stress, providing a measure of the body’s internal response to environmental or physiological stressors during pregnancy [16,17].

During pregnancy, the body undergoes numerous metabolic and immune adaptations to support both maternal and fetal health. These changes can affect the metabolism of drugs, including antimalarial treatments like sulfadoxine-pyrimethamine (SP). Given that lignin-related compounds have been linked to oxidative processes, it is plausible that they could play a role in drug metabolism or stress responses in pregnant women [10,15]. For example, oxidative stress caused by malaria infection or other environmental factors may influence how drugs like SP are processed in the body. If lignin positivity correlates with altered drug metabolism or therapeutic response, it could serve as a useful biomarker to assess not only oxidative stress but also the effectiveness of treatment regimens such as SP during pregnancy.

Moreover, the physiological changes of pregnancy itself, such as alterations in hormone levels, blood circulation, and immune function can impact the body’s response to medications. It is conceivable that lignin, as a byproduct of metabolic processes or as part of the oxidative stress response, could provide insights into how a pregnant woman’s body is processing medications and handling various stressors [18,19]. This could lead to a better understanding of how individual variations in lignin levels might influence therapeutic outcomes, helping to personalize treatment approaches and improve drug efficacy. If lignin proves to be a reliable marker for oxidative stress or drug metabolism in pregnant women, it could play a pivotal role in optimizing malaria prevention strategies and improving maternal and fetal health outcomes [20].

### Drug Adherence and Biomarkers

Drug adherence is a critical factor in determining the effectiveness of any treatment regimen, particularly in maternal health, where non-adherence to prescribed medications can lead to significant maternal and fetal complications [14,15]. In the context of malaria treatment and prevention, adherence to the prescribed regimen is essential for achieving optimal outcomes, as inconsistent use of antimalarial drugs such as sulfadoxine-pyrimethamine (SP) can result in treatment failure, continued malaria transmission, and adverse pregnancy outcomes. Despite the importance of adherence, it is often difficult to accurately assess how well patients follow prescribed treatments, especially in pregnant women, where self-reports of medication use can be unreliable due to recall bias or social desirability [16,17].

Over the years, researchers have sought to identify objective biomarkers that can provide more accurate measures of drug adherence. Biomarkers are measurable indicators of physiological processes or drug exposure that can reflect adherence to prescribed treatments or the body’s response to them [8,10,16]. In maternal health, the use of biomarkers to assess adherence has gained increasing attention, particularly for conditions like malaria, where treatment success is highly dependent on consistent and correct use of medications. Biomarkers can provide insights into whether the drug has been taken as prescribed and can also offer information on the drug’s effectiveness, metabolism, and potential therapeutic response [4,5,18].

In malaria treatment, several biomarkers have been explored to assess adherence to antimalarial drugs [3,4,6]. For example, plasma concentrations of the drug itself or its metabolites can be used to directly measure exposure to the medication. This approach, however, requires frequent blood sampling and sophisticated testing, making it impractical for routine use in resource-limited settings. Other potential biomarkers include indicators of the body’s immune response to malaria, such as changes in specific cytokine levels, or markers of oxidative stress, which may be linked to the body’s response to both the malaria infection and the antimalarial drug [7,9].

One promising avenue for monitoring adherence in malaria treatments is the identification of novel biomarkers that reflect both drug use and the physiological impact of treatment. Lignin, for instance, has the potential to serve as a biomarker for oxidative stress, which could be associated with the body’s response to antimalarial drugs like SP [2,6,8]. By correlating lignin positivity with self-reported drug use or drug concentrations in the body, researchers may be able to develop a more reliable method for assessing adherence and treatment efficacy. If lignin levels are found to reflect the body’s response to SP or other malaria treatments, they could be used to monitor how well a pregnant woman is adhering to the prescribed regimen and whether the drug is having the desired effect on malaria prevention [12,13].

Biomarkers could thus play a pivotal role in improving adherence monitoring and treatment outcomes in maternal health. By providing an objective measure of adherence and therapeutic response, biomarkers can help healthcare providers identify non-adherence early, allowing for timely interventions to improve treatment outcomes [11]. In the context of malaria, using biomarkers to monitor adherence could not only improve the effectiveness of malaria prevention strategies but also ensure better maternal and fetal health outcomes, reducing the risk of complications associated with malaria infection during pregnancy. Furthermore, biomarkers could help tailor treatment regimens to individual needs, optimizing drug efficacy and minimizing potential side effects, thereby enhancing overall treatment success.

### Sulfadoxine-Pyrimethamine (SP) in Malaria Prevention during Pregnancy and the Role of Lignin as a Potential Biomarker for Oxidative Stress

Sulfadoxine-pyrimethamine (SP) is a widely used combination antimalarial drug, essential for the prevention and treatment of malaria, particularly among pregnant women in endemic regions. Malaria during pregnancy presents significant risks to both maternal and fetal health, including maternal anemia, fetal growth restriction, preterm birth, and, in severe cases, maternal and neonatal mortality [13,14]. Therefore, SP is often included as part of intermittent preventive treatment (IPTp) to mitigate these risks and reduce the adverse outcomes associated with malaria.

The drug functions by inhibiting the folate synthesis pathway in malaria parasites, which prevents their replication and subsequently reduces parasite load in the body [16,17].

Malaria during pregnancy remains a substantial public health burden, with estimates suggesting that it accounts for approximately 11% of maternal deaths in malaria-endemic regions [1]. Pregnant women are particularly vulnerable to severe malaria and its associated complications due to physiological changes during pregnancy, which alter immune responses and increase susceptibility to infection. According to a meta-analysis on malaria in pregnancy, it is associated with an increased risk of low birth weight and preterm birth, with a 10-20% higher likelihood of these adverse outcomes compared to women without malaria [19]. Maternal anemia is another significant complication, with SP treatment shown to reduce anemia by 50-60%, significantly improving maternal health in endemic areas.

Intermittent preventive treatment with SP (IPTp-SP) has been demonstrated to be highly effective in reducing both the incidence of malaria and its associated complications. A large-scale randomized controlled trial conducted in sub-Saharan Africa showed that SP administration reduced malaria incidence by up to 70% in pregnant women, as compared to placebo. Additionally, IPTp-SP has been shown to reduce the risk of low birth weight by as much as 40%, further supporting the efficacy of this intervention in improving both maternal and neonatal outcomes [1].

In terms of its mechanism of action, SP works by inhibiting the folate pathway in Plasmodium species, blocking the parasite’s ability to replicate. The efficacy of SP in clearing parasitemia is well-established, with studies showing that SP can reduce parasitemia levels by more than 90% in pregnant women with uncomplicated malaria [3, 10]. This substantial reduction in parasite burden contributes directly to the reduction in malaria-related maternal complications, including anemia and poor fetal outcomes [16].

While SP has proven to be effective in malaria prevention and treatment, the role of lignin, a complex plant polymer, in pregnancy-related oxidative stress and its potential impact on drug metabolism warrants further exploration. Lignin, primarily recognized for its structural role in plants, has been implicated in oxidative stress within the human body. Oxidative stress, defined as an imbalance between reactive oxygen species (ROS) and antioxidants, is a critical factor in various physiological and pathological processes, including immune responses, inflammatory conditions, and drug metabolism [14].

During pregnancy, oxidative stress is often heightened due to increased metabolic demands and changes in the immune system. Research has shown that oxidative stress markers, such as malondialdehyde (MDA), are significantly elevated in pregnant women, particularly those with conditions like pre-eclampsia and gestational diabetes [5,7,9]. Elevated oxidative stress during pregnancy has been linked to alterations in drug metabolism, as it affects the cytochrome P450 enzyme system, which plays a crucial role in the metabolism of many drugs, including sulfadoxine, the active compound in SP. Increased oxidative stress may influence the activity of CYP450 enzymes, potentially altering the pharmacokinetics of SP and other medications, thereby affecting their efficacy and safety in pregnant women [12,13].

The potential relationship between lignin and oxidative stress has not been fully explored in human physiology, but emerging studies suggest that lignin metabolites may reflect changes in oxidative stress levels. Research demonstrated that lignin-derived compounds such as ferulic acid are associated with increased antioxidant activity, potentially indicating that changes in lignin levels could serve as biomarkers for oxidative stress during pregnancy. This is particularly relevant for understanding how drugs like SP are metabolized, as oxidative stress can modulate enzyme activity and influence the absorption, distribution, metabolism, and excretion (ADME) of drugs [14].

Although lignin’s role in human metabolism remains an area for further research, it is plausible that changes in lignin or its metabolites could provide valuable insights into the body’s response to external factors such as drug administration during pregnancy [1,12]. If lignin positivity reflects oxidative stress or immune function changes, it could offer a novel means of understanding how drugs like SP are processed in pregnant women, ultimately influencing treatment outcomes and maternal-fetal health.

## 2. Materials and Methods

### Study Design

This study adopted a longitudinal design to assess the correlation between lignin positivity and self-reported sulfadoxine-pyrimethamine (SP) administration among pregnant women living in malaria-endemic regions. A cross-sectional approach allows the collection of data at a single point in time, providing a snapshot of the relationship between SP use and lignin positivity. The design facilitated the examination of the potential biomarkers of adherence to SP treatment while considering the diverse factors influencing maternal health outcomes.

The study used structured questionnaires on self-reports of SP administration and biomarker analysis to measure lignin positivity. By capturing both quantitative and biochemical data, the study comprehensively assessed adherence and the metabolic impact of SP use during pregnancy. The study received ethical approval from the Joint Ethics Committee of the University of Ibadan and the University College Hospital, Ibadan (UI/UCH/EC/24/0305).

### Population

The study focused on all pregnant women aged 18-45 years living in malaria-endemic regions, as this group is most at risk for malaria and its complications during pregnancy.

### Sample size and Sampling Techniques

The samples were recruited from Adeoyo Maternity Teaching Hospital, Yemetu, Ibadan. All pregnant women attending booking and routine antenatal care clinics who consented to participate in the study were included as subjects. The Adeoyo Maternity Hospital (AMH), Ibadan was found suitable for the study because it provides comprehensive malaria prevention services, including regular antenatal care, health education, and distribution of insecticide-treated nets (ITNs). Additionally, the hospital serves a large and diverse population of pregnant women, making it an ideal setting for studying malaria prevention practices during pregnancy.

### Inclusion Criteria

i. Pregnant women in the second or third trimester at-least at gestation age of 16weeks or above
ii. Women who have received SP for malaria prevention during pregnancy.
iii. Women who have access to healthcare services providing SP for malaria prophylaxis.
iv. Consent to study protocol.

### Exclusion Criteria

a. Women with pre-existing conditions that could interfere with SP metabolism or affect lignin levels, such as renal disease, liver disease, or autoimmune conditions. These conditions may alter the metabolism of SP or impact oxidative stress levels.
b. Women with non-pregnancy-related health conditions such as HIV or cancer that may interfere with the assessment of drug adherence or lignin metabolism.
c. History suggestive of serious adverse reaction to sulpha drugs.

### Sample Size Determination

The minimum number of subjects studied was obtained using the sample size formula (Charan and Biswas, 2013).

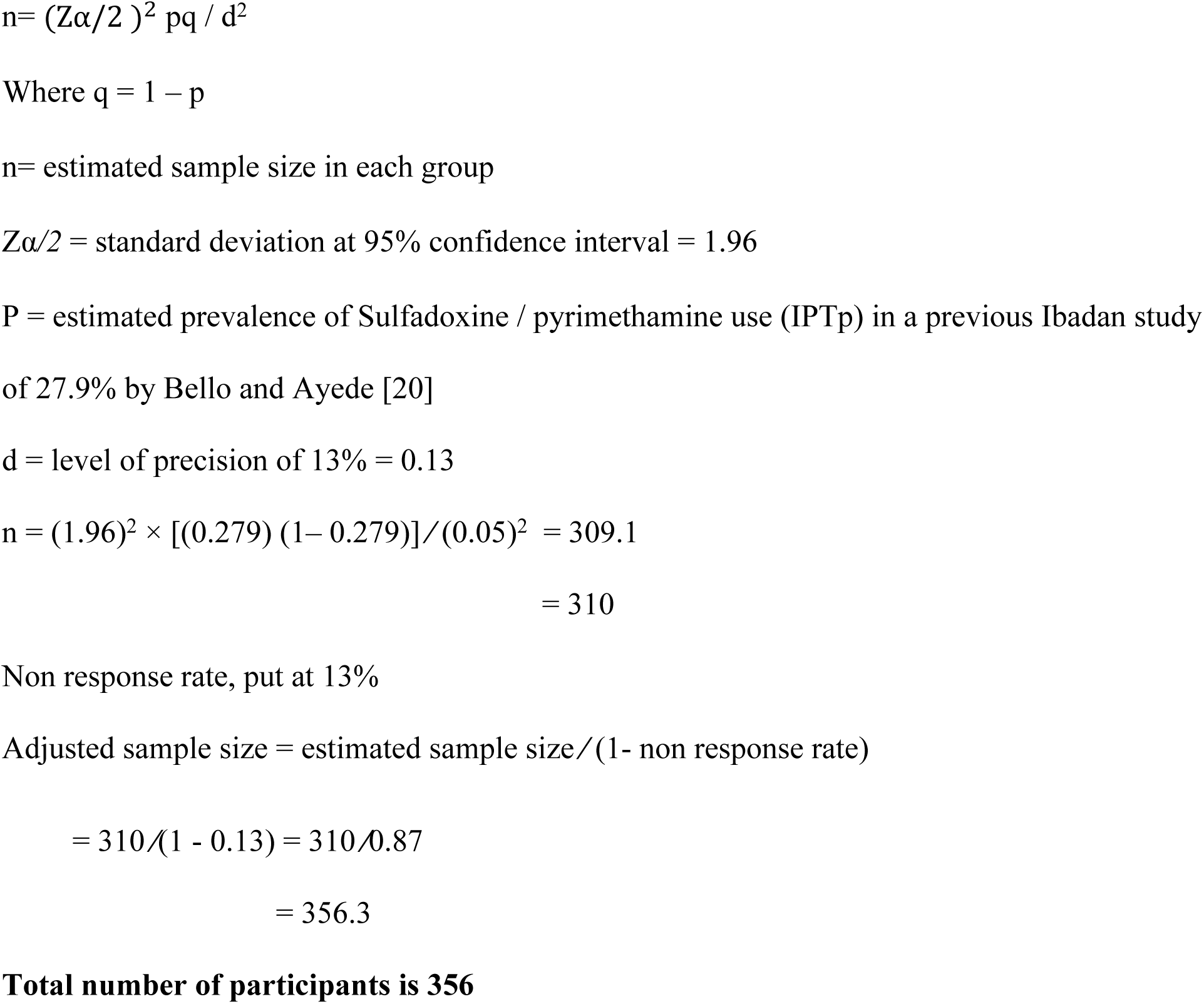

### Data Collection

Data on SP administration were collected using structured researcher’s self-developed questionnaire where participants reported on the dosage, frequency and any missed doses of SP taken. The questionnaire included questions about the woman’s health status, and any challenges faced in accessing or taking SP as prescribed.

Urine samples were collected from participants for biochemical assays to measure lignin markers using a drop of 10% hydrogen chloride to a newsprint containing, which to indicate oxidative stress or the metabolic response to SP administration. Lignin positivity was determined by analyzing specific oxidative stress markers associated with lignin and comparing them against established reference levels of the selected pregnant women.

### Statistical Analysis

Descriptive statistics with the use of mean and standard deviation was employed to summarize the demographic characteristics of the sample and the prevalence of lignin positivity. This will include frequency distributions for categorical variables (e.g., age ranges, socio-economic status, history of malaria) and measures of central tendency (mean, median) for continuous variables (e.g., age, number of pregnancies, gestational age).

To assess the relationship between lignin positivity and self-reported SP administration, Pearson Product Moment Correlation (PPMC) was used for normally distributed variables. This analysis helped determine if there is a statistically significant relationship between lignin levels and SP adherence.

### Ethical Considerations

Ethical approval was sought from the Joint Ethics Committee of the University of Ibadan and the University College Hospital, Ibadan. Participants provided informed consent prior to participation, ensuring they are fully aware of the study’s aims, procedures, and any potential risks. The confidentiality of all participants were maintained throughout the study, with all data anonymised and securely stored.

## 3. Results

In this study, only 356 pregnant women were targeted but one (1) data was missing due to withdrawal by 1 participant, hence only 355 data were processed, as malaria chemoprophylaxis with IPTp-SP were analyzed to understand various aspects regarding their socio-demographic characteristics and their backgrounds as being recruited between July 2024 and November 2024.

The socio-demographic characteristics of 355 pregnant women on malaria chemoprophylaxis who self-reported administration of Sulfadoxine-Pyrimethamine (SRA-SP) were as follows: The average age of the participants was 29.9 years (SD = 5.6). The age distribution revealed that 4.5% were under 20 years, 44.8% were between 20 and 29 years, 45.4% were aged 30 to 39 years, and 5.3% were 40 years or older. The age distribution showed that the majority of women (90.2%) were aged between 21 and 39 years, with 44.8% in the 20-29 age group and 45.4% in the 30-39 age group. This is consistent with the demographic trend in many populations where childbearing tends to peak in the third decade of life. The relatively low percentage (4.5%) of women under 20 years old suggests that younger women may have lower access to healthcare resources or might be less likely to participate in antenatal care and malaria chemoprophylaxis. The 6.3% of women aged 40 years and above aligns with the observation that women in older age groups tend to have fewer pregnancies, as fertility rates typically decline with age.

Regarding marital status, the majority (97.5%) were married, while 2.5% were single. A significant proportion of participants (97.5%) were married, which reflects societal norms and expectations in many regions, particularly in sub-Saharan Africa, where marriage is a prominent institution in reproductive health practices. The very low percentage of single women (2.5%) is consistent with traditional family structures where marriage precedes childbearing. The marital status distribution also likely reflects the higher participation of married women in maternal healthcare programmes such as malaria chemoprophylaxis.

Family type was predominantly monogamous, accounting for 89.3%, with 7.6% in polygamous households and 0.6% offering no response. The majority of women (89.3%) were in monogamous marriages, while 7.6% were in polygamous households. This result can be explained by the cultural and societal norms prevalent in the study region, where monogamy is more common but polygamy still exists in certain areas, especially in specific ethnic or social groups. The relatively low number of non-responses (3.1%) suggests that family type is a well-recognised and easily identifiable characteristic in the population of selected pregnant women attending University College Hospital, Ibadan.

In terms of education, 50.3% had completed tertiary education, 37.6% had secondary education, 5.1% had primary education, and 0.8% had no formal education. Only 0.3% did not provide an educational response. The highest educational attainment observed was tertiary education, with 50.3% of participants reporting having completed this level. This is indicative of the increasing access to higher education in many parts of the world, especially in urban or more developed regions. It also reflects the importance of education in empowering women, particularly in health-seeking behaviours such as attending antenatal care and using malaria chemoprophylaxis. A significant proportion (37.6%) had completed secondary education, which suggests that while many women have access to basic education, opportunities for further education may still be limited for some. The low number of women with no formal education (0.8%) highlights improvements in education over time in the region.

Occupation-wise, 88.7% were employed, 10.4% were unemployed, and 0.8% did not provide a response. The majority (88.7%) of women were employed, which suggests that many in this sample may have access to employment opportunities, possibly contributing to better socio-economic status and healthcare access. The 10.4% unemployment rate reflects the broader socio-economic challenges faced by certain sections of the population, which may include rural or less-developed areas where employment opportunities are less available. The negligible number of non-responses (0.8%) indicates that employment status is a well-documented attribute in the survey.

Ethnically, 97.2% were Yoruba, followed by 1.7% Igbo, 0.6% Hausa, and 0.3% from other groups, with 0.3% offering no ethnic response. The overwhelming majority of participants (97.2%) were Yoruba, with smaller percentages of Igbo (1.7%), Hausa (0.6%), and other ethnicities (0.3%). This reflects the ethnic composition of the region where the study was conducted. The Yoruba ethnic group is prominent in southwestern Nigeria, which explains the high percentage, while the smaller percentages of other ethnicities reflect the diversity that exists within the population.

On obstetric history, the mean gravida (number of pregnancies) was 2.4 (SD = 1.4, range 1-9). The mean number of abortions was 0.4 (SD = 0.7, range 0-3), and the mean number of living children was 1.4 (SD = 1.3, range 1-8). The mean gravida (2.4) indicates that the women in this cohort generally had a moderate number of pregnancies. This is typical of women in this age range and reflects the general fertility pattern in sub-Saharan Africa. The mean number of abortions (0.4) is relatively low, indicating that most women had not experienced multiple miscarriages or elective abortions. The average number of living children (1.4) is lower than the mean gravida, suggesting that some women may have had pregnancies that did not result in live births, possibly due to stillbirths, miscarriages, or other adverse outcomes.

### Testing of Hypothesis

H_o_: There is no statistically significant correlation between lignin positivity and the self-reported administration of sulfadoxine-pyrimethamine (SP) among pregnant women.

The table 2 below presents the results of the lignin test over three time points: Day 0, Day 7, and Day 28. On Day 0, the test showed that 58.6% of the participants had a negative result, while 41.4% had a positive result. The mean lignin test score was 0.41 with a standard deviation of 0.493, and the p-value indicated no significant difference (p > 0.05).

**Table 1:**
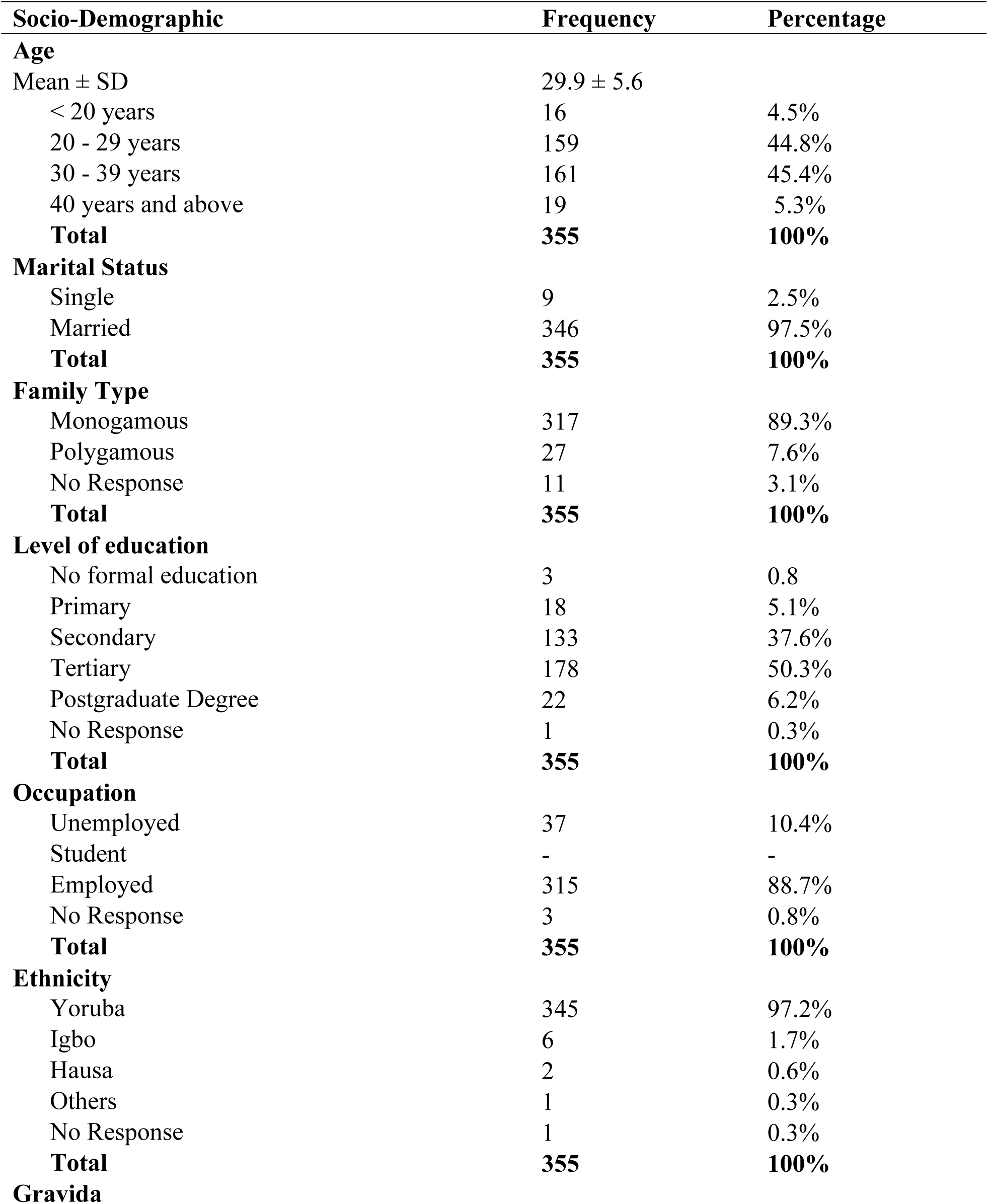
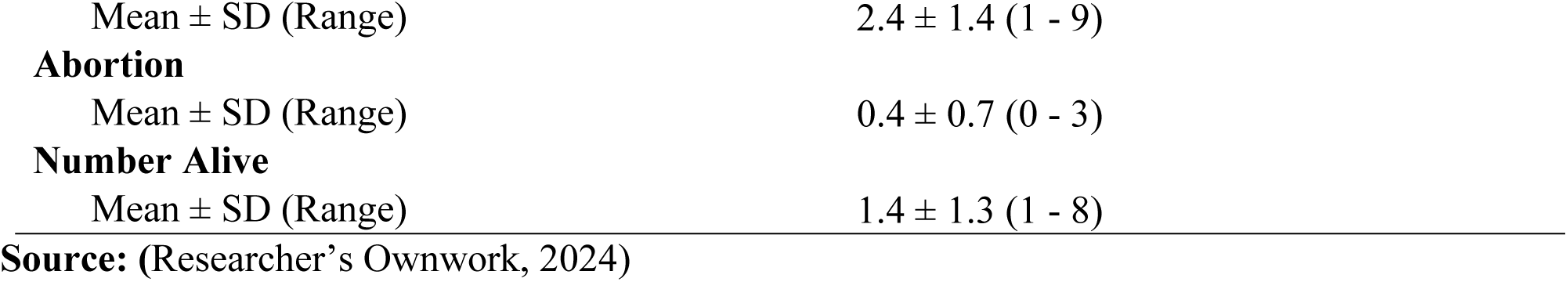
Socio Demographic Characteristics of 355 pregnant women on malaria chemoprophylaxis with Self-Reported Administration of Sulfadoxine-Pyrimethamine (SRA-SP)

| **Socio-Demographic** | **Frequency** | **Percentage** |
| --- | --- | --- |
| **Age** |  |  |
| Mean ± SD | 29.9 ± 5.6 | |
| < 20 years | 16 | 4.5% |
| 20 - 29 years | 159 | 44.8% |
| 30 - 39 years | 161 | 45.4% |
| 40 years and above | 19 | 5.3% |
| **Total** | **355** | **100%** |
| **Marital Status** |  |  |
| Single | 9 | 2.5% |
| Married | 346 | 97.5% |
| **Total** | **355** | **100%** |
| **Family Type** |  |  |
| Monogamous | 317 | 89.3% |
| Polygamous | 27 | 7.6% |
| No Response | 11 | 3.1% |
| **Total** | **355** | **100%** |
| **Level of education** |  |  |
| No formal education | 3 | 0.8 |
| Primary | 18 | 5.1% |
| Secondary | 133 | 37.6% |
| Tertiary | 178 | 50.3% |
| Postgraduate Degree | 22 | 6.2% |
| No Response | 1 | 0.3% |
| **Total** | **355** | **100%** |
| **Occupation** |  |  |
| Unemployed | 37 | 10.4% |
| Student | - | - |
| Employed | 315 | 88.7% |
| No Response | 3 | 0.8% |
| **Total** | **355** | **100%** |
| **Ethnicity** |  |  |
| Yoruba | 345 | 97.2% |
| Igbo | 6 | 1.7% |
| Hausa | 2 | 0.6% |
| Others | 1 | 0.3% |
| No Response | 1 | 0.3% |
| **Total** | **355** | **100%** |
| **Gravida** |  |  |

|  |  |
| --- | --- |
| Mean ± SD (Range) | 2.4 ± 1.4 (1 - 9) |
| **Abortion** |  |
| Mean ± SD (Range) | 0.4 ± 0.7 (0 - 3) |
| **Number Alive** |  |
| Mean ± SD (Range) | 1.4 ± 1.3 (1 - 8) |

**Source:** (Researcher's Ownwork, 2024)

**Table 2:** Pearson Product Moment Correlation (PPMC) (r) statistics showing relationship between lignin positivity and the self-reported administration of sulfadoxine-pyrimethamine (SP) among pregnant women at Day 0.

| Lignin Test Result | Lignin Test Result |  | Mean | Std. Dev. | r-Cal. | p-value | Decision |
| --- | --- | --- | --- | --- | --- | --- | --- |
|  | Day 0 |  |  |  |  |  |  |
|  | Frequency | Percentages % |  |  |  |  |  |
| **Day 0** |  |  | 0.41 | 0.493 | 1 | . | **Sig.** |
| Negative | 208 | 58.6 |  |  |  |  |  |
| Positive | 147 | 41.4 |  |  |  |  |  |
| **Total** | **355** | **100.0** |  |  |  |  |  |

\*Significant at <0.05 alpha level. **Source:** (Researcher's Ownwork, 2024)

By Day 7, a significant change was observed. The mean score increased to 0.92 (SD = 0.279), with 91.3% of participants testing positive, 8.5% testing negative, and 0.3% providing an invalid result.

The p-value of 0.000 confirmed the result was statistically significant, indicating a clear shift towards more positive test outcomes compared to Day 0.

On Day 28, the test results remained predominantly positive, with 93.5% testing positive, 3.9% negative, and 2.5% invalid. The mean score reached 0.96 (SD = 0.197), with a p-value of 0.007, further confirming the statistical significance of the results. This suggests a sustained increase in positive lignin test results from Day 0 through Day 28.

The null hypothesis (H_0_) posited that there was statistical significant correlation between lignin positivity and the self-reported administration of sulfadoxine-pyrimethamine (SP) among pregnant women. The alternative hypothesis (H_1_) suggested that there was a statistically significant correlation.

The results from the lignin test conducted at three time points (Day 0, Day 7, and Day 28) provide evidence that supports the rejection of the null hypothesis. On Day 0, the proportion of positive lignin test results was 41.4%, with a mean score of 0.41, indicating no significant change (p > 0.05). However, by Day 7, the percentage of positive results increased sharply to 91.3%, with a significant mean score of 0.92 (p = 0.000). This substantial shift in test outcomes suggests that the administration of sulfadoxine-pyrimethamine may have had an impact on lignin positivity.

By Day 28, the positivity rate continued to rise to 93.5%, with a mean score of 0.96 and a significant p-value of 0.007. The sustained increase in positive lignin results over the 28-day period, coupled with the statistically significant p-values at both Day 7 and Day 28, indicates a correlation between lignin positivity and the self-reported use of SP (See table 2 to 4 below).

Thus, the results provide compelling evidence that there was a statistical significant correlation between lignin positivity and the administration of sulfadoxine-pyrimethamine, supporting the rejection of the null hypothesis and confirming the validity of the alternative hypothesis.

This is an indication that the level of adherence to the self-reported administration of SP was high, with a statistically significant correlation between SP use and the increase in positive lignin test results. The adherence of pregnant women to the self-reported administration of sulfadoxine-pyrimethamine (SP) has been confirmed by the lignin test that over the 28-day period of SP usage. As illustrated on table 3 below, the significant increase in positive lignin test results from Day 0 to Day 28, with 41.4% of participants testing positive on Day 0, 91.3% on Day 7, and 93.5% on Day 28, indicates a high level of adherence to the self-reported administration of SP. This is an indication that the majority of pregnant women who self-reported SP administration did indeed follow through with the treatment, leading to a clear shift in the test outcomes. This progression continued through Day 0 to 28, further reinforcing the idea that most women adhered to the prescribed malaria chemoprophylaxis regimen.

**Table 3:**
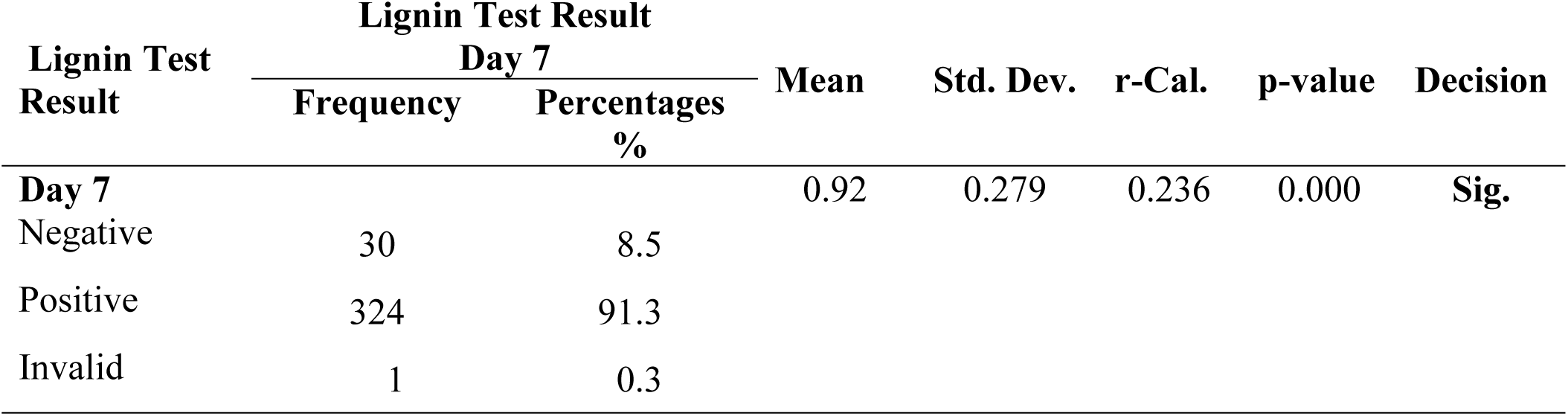
Pearson Product Moment Correlation (PPMC) (r) statistics showing relationship between lignin positivity and the self-reported administration of sulfadoxine-pyrimethamine (SP) among pregnant women at Day 7.

| Lignin Test Result | Lignin Test Result |  | Mean | Std. Dev. | r-Cal. | p-value | Decision |
| --- | --- | --- | --- | --- | --- | --- | --- |
|  | Day 7 |  |  |  |  |  |  |
|  | Frequency | Percentages % |  |  |  |  |  |
| Day 7 |  |  | 0.92 | 0.279 | 0.236 | 0.000 | Sig. |
| Negative | 30 | 8.5 |  |  |  |  |  |
| Positive | 324 | 91.3 |  |  |  |  |  |
| Invalid | 1 | 0.3 |  |  |  |  |  |

By Day 7, a significant change was observed. The mean score increased to 0.92 (SD = 0.279), with 91.3% of participants testing positive, 8.5% testing negative, and 0.3% providing an invalid result. The p-value of 0.000 confirmed the result was statistically significant, indicating a clear shift
towards more positive test outcomes compared to Day 0.

**Table 4:**
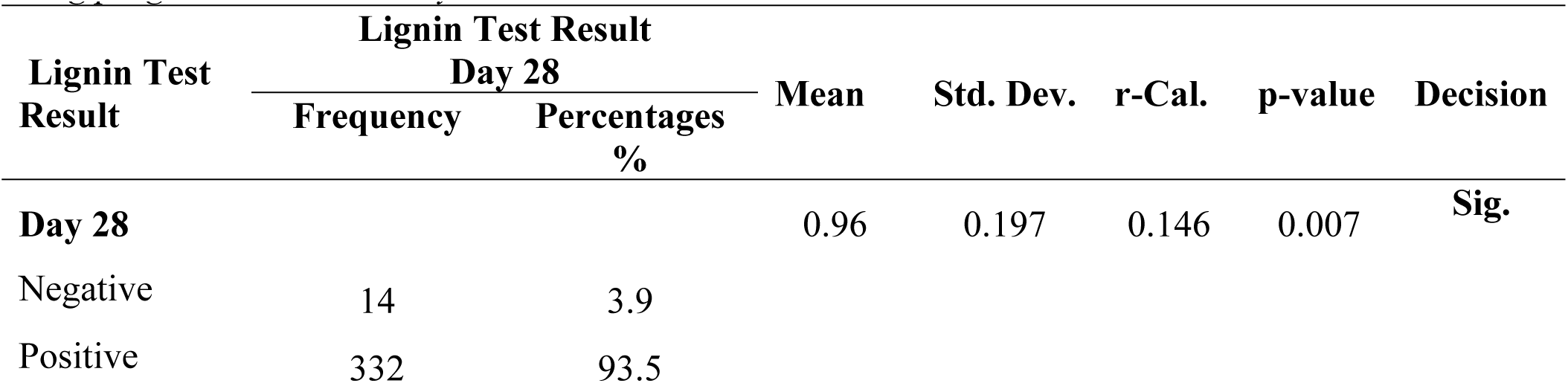
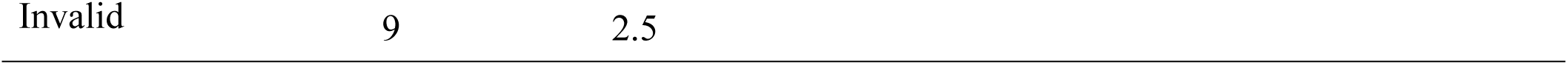
Pearson Product Moment Correlation (PPMC) (r) statistics showing relationship between lignin positivity and the self-reported administration of sulfadoxine-pyrimethamine (SP) among pregnant women at Day 28.

## 4. Discussion

The findings of this study provide strong evidence to reject the null hypothesis (H_0_), which suggested statistical significant correlation between lignin positivity and the self-reported administration of sulfadoxine-pyrimethamine (SP) among pregnant women. The results from the lignin test at multiple time points (Day 0, Day 7, and Day 28) clearly demonstrate a statistically significant increase in lignin positivity over the course of the study period, with a marked rise in positive test results correlating with the self-reported administration of SP. This increase in lignin positivity suggests that SP administration plays a role in promoting higher oxidative stress, potentially as part of the body’s therapeutic response to the drug.

These findings are consistent with previous research by Chico et al. [8] highlighting the effectiveness of sulfadoxine-pyrimethamine in malaria prevention among pregnant women. Additionally, study by Green et al. [15] have shown that the use of SP significantly reduces the incidence of malaria and its associated adverse outcomes, such as maternal anaemia, low birth weight, and preterm delivery. Moreover, Gascoigne et al. [3] also found that the association between lignin positivity and SP administration underscores the potential of lignin as a biomarker for assessing adherence to malaria chemoprophylaxis, an essential factor in ensuring the success of preventive interventions.

The significant increase in lignin positivity observed in this study also aligns with the broader efforts to assess and improve adherence to intermittent preventive treatment (IPTp) in pregnancy [1]. Given credence to the study by Baraka et al. [4], it was stated that high adherence to SP treatment is crucial in regions where malaria remains endemic, and improving treatment compliance can contribute significantly to the reduction of malaria-related maternal and neonatal morbidity and mortality. The study’s findings suggest that self-reported data, when coupled with biochemical markers like lignin, may offer a reliable means of monitoring adherence to SP treatment. This study contributes to the growing body of evidence supporting the use of SP for malaria prevention in pregnancy and highlights the importance of adherence in determining the efficacy of such interventions.

## 5. Conclusion

The study investigated the correlation between lignin positivity and the self-reported administration of sulfadoxine-pyrimethamine (SP) in pregnant women in Yemetu, Ibadan. It was found that there existed strong evidence supporting a statistically significant correlation between lignin positivity and the self-reported administration of sulfadoxine-pyrimethamine (SP) among pregnant women. The results from the lignin test at three time points indicate a clear increase in positive test outcomes over time, which was statistically significant. This progression suggests that the administration of SP had a measurable impact on lignin positivity.

The findings indicate a high level of adherence to the self-reported administration of SP, with most participants consistently following the prescribed malaria chemoprophylaxis regimen among pregnant women attending Adeoyo Maternity Teaching Hospital, Yemetu, Ibadan. The sustained increase in positive lignin test results throughout the study period further reinforces the idea that the majority of pregnant women adhered to the treatment. Seamlessly, the study reflected on the effectiveness of SP in malaria prevention and highlights the high adherence levels among pregnant women.

## Data Availability

All relevant data underlying the findings of this study titled “Correlating Lignin Positivity with Self-Reported Administration of Sulfadoxine-Pyrimethamine (SRA-SP) Among Pregnant Women” are fully available without restriction. The de-identified dataset used to support the findings, including participant responses to the self-structured questionnaire, lignin positivity results across Day 0, Day 7, and Day 28 intervals, and associated demographic information too has been deposited in a public open-access repository. Researchers seeking additional information may also contact the corresponding author, Dr Dhikrullah A. Adebayo, at. There are no legal or ethical restrictions preventing public sharing of the anonymised data. All participants provided informed consent and data were handled in accordance with ethical approvals obtained from the appropriate institutional review boards.

## Acknowledgments

The authors extend their sincere appreciation to the management and antenatal care staff of Adeoyo Maternity Teaching Hospital, Yemetu, Ibadan, for their unwavering support during data collection. We also acknowledge the pregnant women who voluntarily participated in this study, for their cooperation, patience, and commitment throughout the follow-up intervals.

Special thanks go to the laboratory technologists and data collection assistants whose diligence and professionalism contributed immensely to the success of the biochemical analyses and interviews conducted.

---

**Figure 1.**
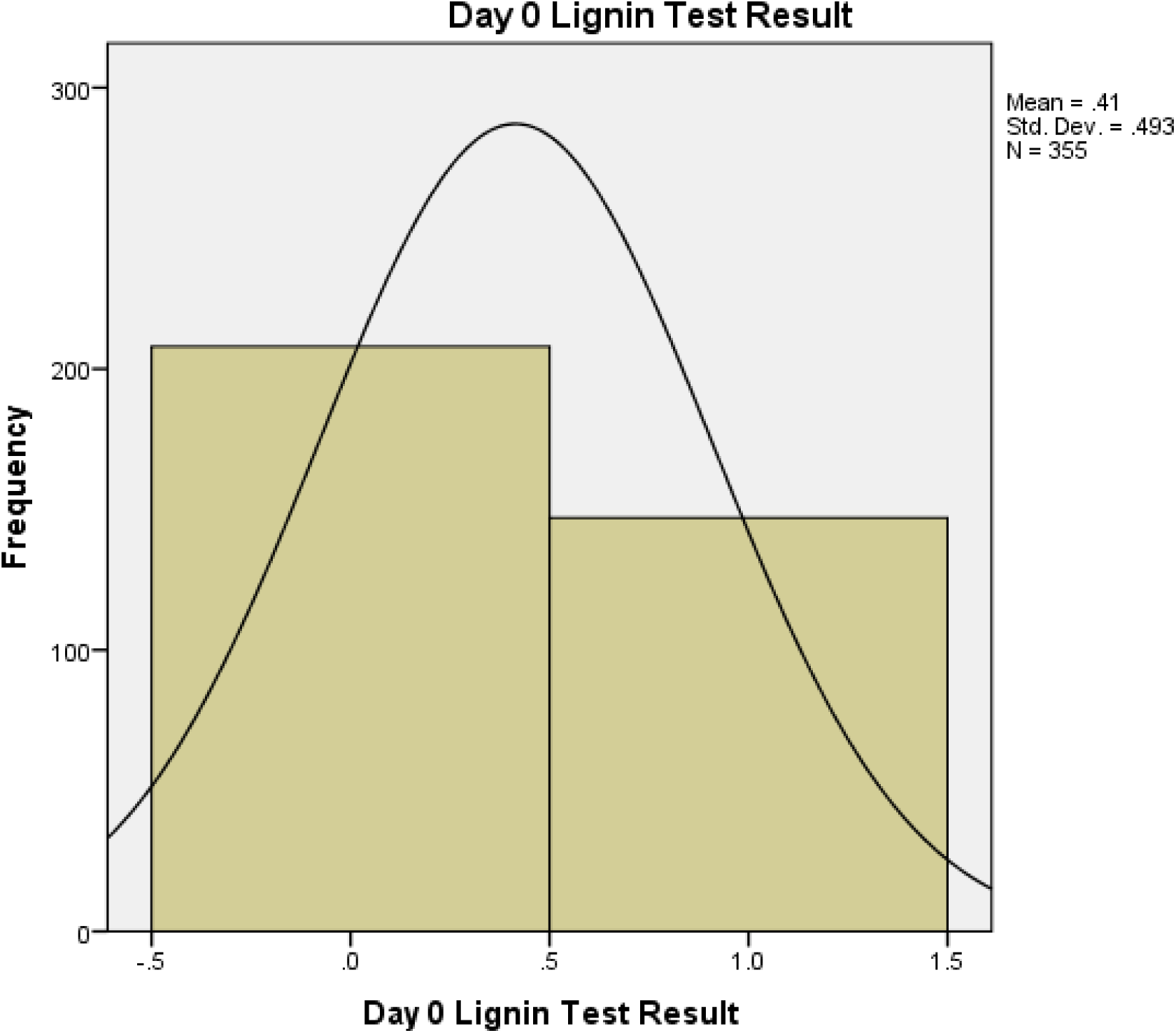
Descriptive Illustration of Lignin Test Result at Day 0 (Researcher’s Ownwork, 2024)

**Figure 2.**
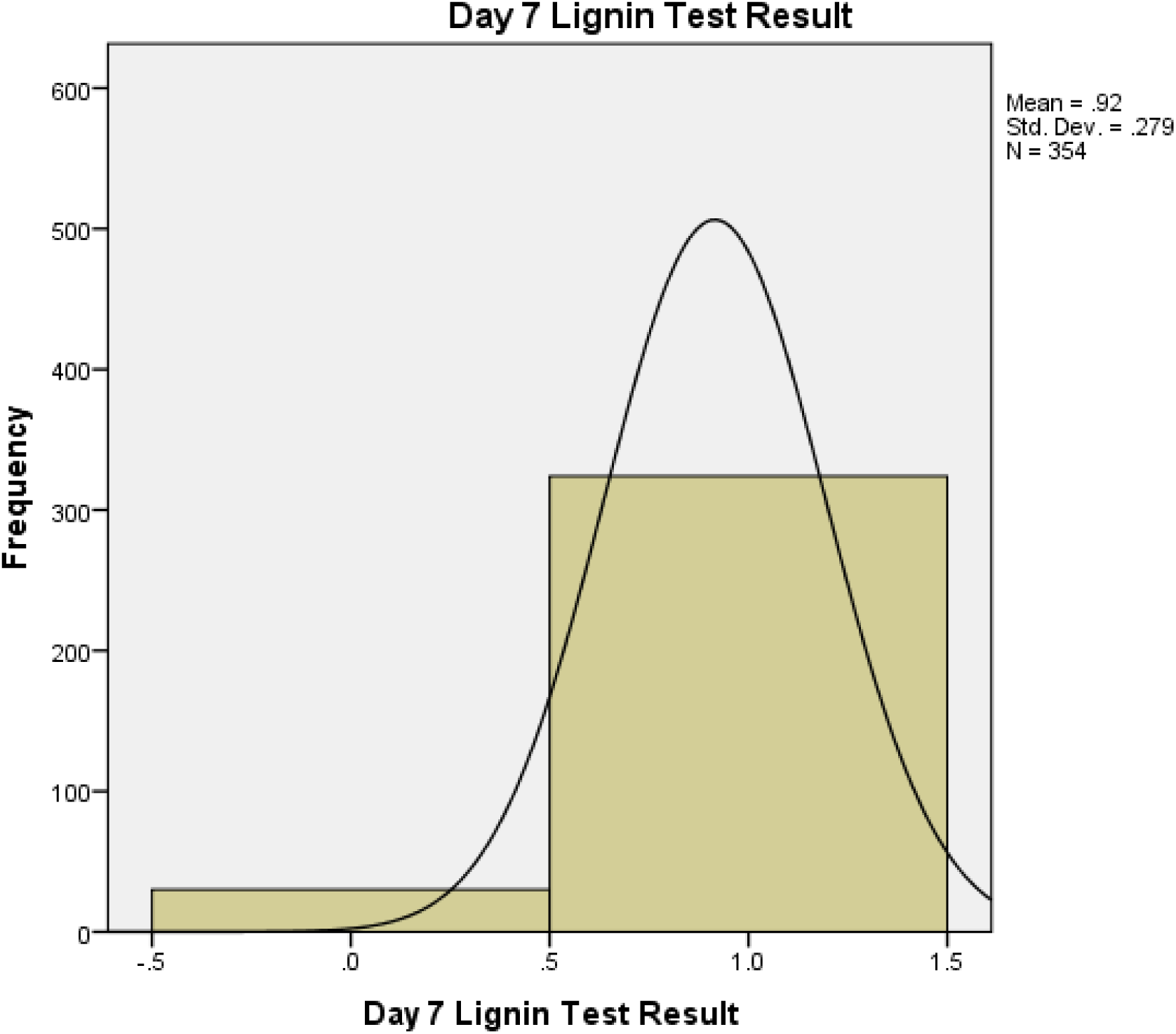
Descriptive Illustration of Lignin Test Result at Day 7(Researcher’s Ownwork, 2024)

**Figure 3.**
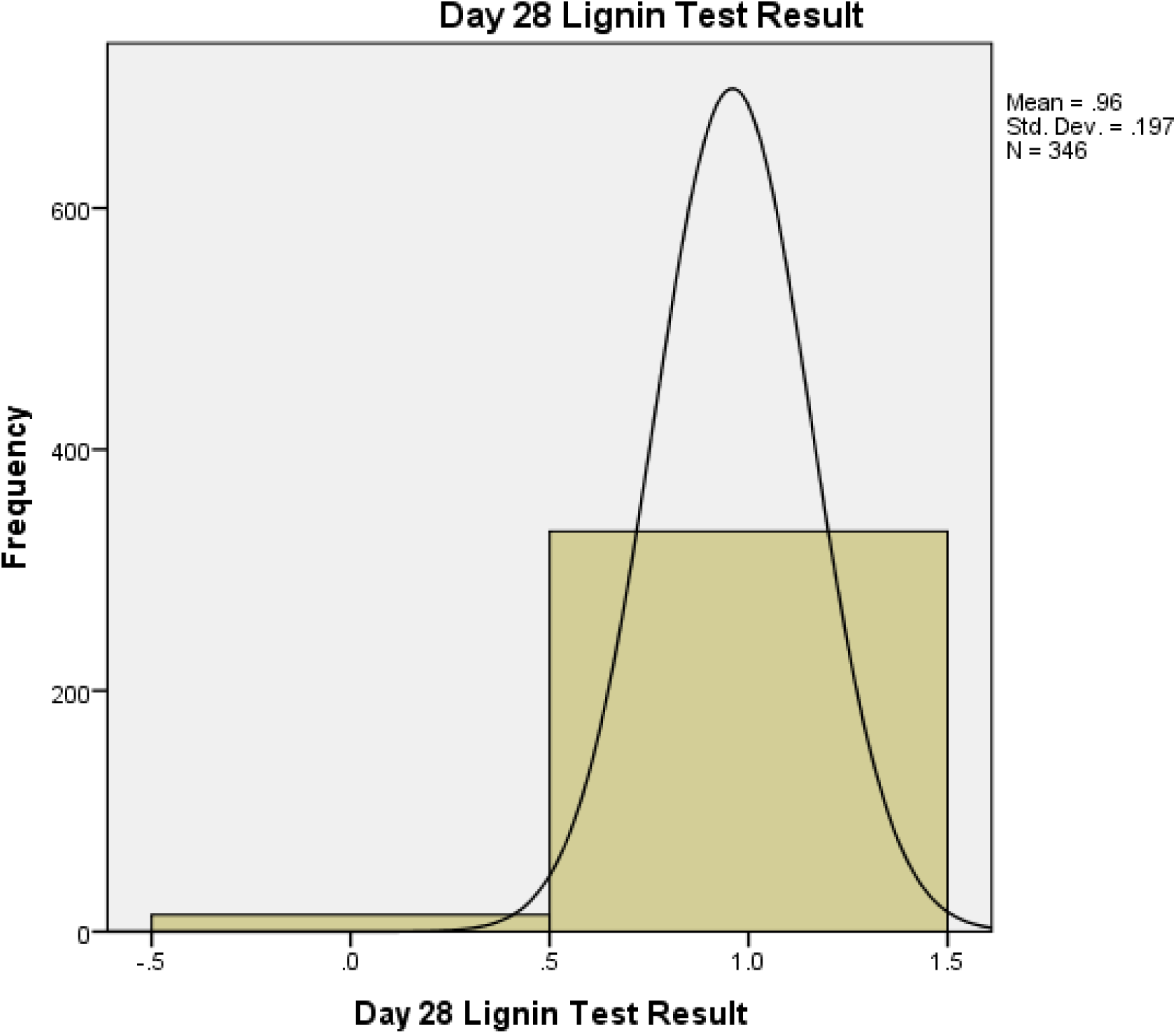
Descriptive Illustration of Lignin Test Result at Day 28 (Researcher’s Ownwork, 2024)

## References

1. World Health Organization. World Malaria Report 2023. World Health Organization, 2023. WHO. Policy Brief for the Implementation of Intermittent Preventive Treatment of Malaria in Pregnancy. World Health Organization, 2014.

2 World Malaria Report. 20 Years of Global Progress and Challenges. World Health Organization, 2020. Licence: CC BY-NC-SA 3.0 IGO. Accessed 31 Jan. 2025.

3 Gascoigne, D. A., Minhaj, M. M., and Aksenov, D. P. “Neonatal Anesthesia and Oxidative Stress.” Antioxidants, vol. 11, no. 4, 2022, p. 787.

4 Baraka, V., Ishengoma, D.S., Fransis, F., Minja, D.T.R., Madebe, R.A., Ngatunga, D., et al. “High-level Plasmodium falciparum Sulfadoxine-pyrimethamine Resistance with the Concomitant Occurrence of Septuple Haplotype in Tanzania.” Malaria Journal, vol. 14, 2015, p. 439.

5 Bhatt, S., Weiss, D.J., Cameron, E., Bisanzio, D., Mappin, B., Dalrymple, U. “The Effect of Malaria Control on Plasmodium falciparum in Africa Between 2000 and 2015.” Nature, vol. 526, no. 7572, 2015, pp. 207–211.

6 Bello, F.A. and Ayede, A.I. “Prevalence of Malaria Parasitaemia and the Use of Malaria Prevention Measures in Pregnant Women in Ibadan, Nigeria.” Annals of Ibadan Postgraduate Medicine, vol. 17, no. 2, 2019, pp. 124–129.

7 Bouyou-akotet, M.K., Ionete-collard, D.E., Mabika, M., Kendjo, E., Matsiegui, P., Mavoungou, E., and Kombila, M. “Prevalence of Plasmodium falciparum Infection in Pregnant Women in Gabon.” Malaria Journal, vol. 2, no. 18, 2003, pp. 1–7.

8 Chico, R.M., Chaponda, E.B., Ariti, C., and Chandramohan, D. “Sulfadoxine-pyrimethamine Exhibits Dose-Response Protection Against Adverse Birth Outcomes Related to Malaria and Sexually Transmitted and Reproductive Tract Infections.” Clinical Infectious Diseases, vol. 64, no. 8, 2017, pp. 1043–1051.

9 Charan, J., and Biswas, T. “How to Calculate Sample Size for Different Study Designs in Medical Research?” Indian Journal of Psychological Medicine, vol. 35, no. 2, 2013, pp. 121–126.

10 Dawaki, S., Mekhlafi, H.M., Al Ithoi, I., Ibrahim, J., Atroosh, W.M., Abdulsalam, A.M., Lau, Y.L. “Is Nigeria Winning the Battle Against Malaria? Prevalence, Risk Factors, and KAP Assessment Among Hausa Communities in Kano State.” Malaria Journal, vol. 15, no. 351, 2016, pp. 1–14.

11 De Almeida Filho, J., and De Souza, J.M. “A Simple Urine Test for Sulfonamides.” Bulletin of the World Health Organization, vol. 61, no. 1, 1983, pp. 167–168.

12 De Kock, M., Tarning, J., Workman, L., Nyunt, M.M., Adam, I., Barnes, K.I., et al. “Pharmacokinetics of Sulfadoxine and Pyrimethamine for Intermittent Preventive Treatment of Malaria During Pregnancy and After Delivery.” CPT Pharmacometrics Systems Pharmacology, vol. 6, no. 6, 2017, pp. 430–438.

13 Drakeley, C., Abdulla, S., Agnandji, S.T., Fernandes, J.F., Kremsner, P., Lell, B., et al. “Longitudinal Estimation of Plasmodium falciparum Prevalence in Relation to Malaria Prevention Measures in Six Sub-Saharan African Countries.” Malaria Journal, vol. 16, no. 1, 2017, pp. 1–15.

14 Elbadry, M.A., Tagliamonte, M.S., Raccurt, C.P., Lemoine, J.F., Existe, A., Boncy, J., et al. “Submicroscopic Malaria Infections in Pregnant Women From Six Departments in Haiti.” Tropical Medicine and International Health, vol. 22, no. 9, 2017, pp. 1030–1036.

15 Green, M.D., van Eijk, A.M., ter Kuile, F.O., Ayisi, J.G., Parise, M.E., Kager, P.A., et al. “Pharmacokinetics of Sulfadoxine-Pyrimethamine in HIV-Infected Pregnant Women.” Tropical Medicine and International Health, vol. 22, no. 10, 2017, pp. 1201–1209.

16 Mikomangwa, W.P., Oms, M., Aklillu, E., Kamuhabwa, A.A.R. “Adverse Birth Outcomes Among Mothers Who Received Intermittent Preventive Treatment with Sulfadoxine-Pyrimethamine in a Low Malaria Transmission Region.” BMC Pregnancy and Childbirth, vol. 19, 2019, p. 236.

17 Nkumama, I.N., O’Meara, W.P., and Osier, F.H.A. “Changes in Malaria Epidemiology in Africa and New Challenges for Elimination.” Trends in Parasitology, vol. 33, no. 2, 2017, pp. 128–140.

18 Omer, S.A., Noureldein, A.N., Eisa, H., Abdelrahim, M., Idress, H.E., Abdelrazig, A.M., et al. “Impact of Submicroscopic Plasmodium falciparum Parasitaemia on Maternal Anaemia and Low Birth Weight in Blue Nile State.” Sudan Journal of Tropical Medicine, 2019, p. 3162378.

19 Peters, G.O., and Naidoo, M. “Factors Influencing the Use of Intermittent Preventive Treatment of Pregnant Women Seeking Care at Primary Healthcare Facilities in the Healthcare System.” African Journal of Primary Health Care and Family Medicine, vol. 12, no. 1, 2020, pp. 1–8.

20 Bello, F. A., and Ayede, A. I. “Prevalence of Malaria Parasitaemia and the Use of Malaria Prevention Measures in Pregnant Women in Ibadan, Nigeria.” Annals of Ibadan Postgraduate Medicine, vol 17, no. 2, 2019, pp. 124–129.

21 Roman, E., Ngindu, A., Orji, B., Zoungrana, J., Robbins, S., and Brieger, W. “Evolution of Malaria in Pregnancy Control: Jhpiego’s 10-Year Contribution.” International Journal of Gynecology and Obstetrics, vol. 130, S2, 2015, pp. S62–S67.

22 Adebayo, D.A. Assessment of Adherence to Intermittent Preventive Treatment in Pregnancy with Sulfadoxine-Pyrimethamine (IPTp-SP) Using Lignin Test. Unpublished dissertation, Department of Pharmacology and Therapeutics, Faculty of Basic Sciences, University of Ibadan, 2021, pp. 1–51.

